# Past and present of registered nurse in China--descriptive analysis of longitudinal national statistics of registered nurse

**DOI:** 10.1101/2024.09.10.24313443

**Authors:** Lihong Yin, Liwen Qiu, Guozhou Zhang

**Affiliations:** Center for Disease Control and Prevention (CDC) in Huangpu District, Shanghai, China, 200032; The Fourth Rehabilitation Hospital of Shanghai, China, 200040

**Keywords:** registered nurse, national statistics, longitudinal data, descriptive analysis

## Abstract

**Background:** China is now in the midst of period of large population aging, which coincides with a global nurse shortage. What added to this problem is the high turnover rate among nurses.

**Methods:** The longitudinal national statistics of registered nurses were analyzed in order to provide a broad background to understand and interpret the high turnover rate among nurses properly.

**Results:** A great number of beds, predominantly hospital-based beds, has been hurriedly prepared for potential inpatients in past two decades, and accordingly the increase of medical professionals especially registered nurses. The workload discrepancies of registered nurses among three tiers of hospitals may come from different sources.

**Conclusion:** This discrepancy is helpful in understanding of high nurse burnout and turnover rate though it seems as inherent systematic problems. A trend of high quality medical service attracting more inpatients has shown, this may cause concerns rather than an satisfied answer, were this trend continued.

## 1 Introduction

The global nursing workforce reached 29.1 millions in 2020, accounting for 59% of all health professionals, and had become the leading health workforce ^1,2^. From 2020 to 2030, the estimated health workforce shortage will shrink significantly from 15.37 million to 10.23 million (reduced by 33%), among them the nursing personnel shortage decreased from 7.07 million (46%) to 4.50 million(44%)^1^. It seems the gap of nurse shortage is closing. This projection, however, was largely based upon the data (74.1%) collected before the coronavirus disease 2019 (COVID-19) pandemic (outbreak characterized from March 11, 2020 on)^1,3^. The pandemic that put heavy burden on the already vulnerable health system and stretched weary medical workforce greatly may accelerate the outflow of nurse population. A pooled analysis (18 studies) of prevalence of turnover intention among intensive cure nurses is 28%(95% CI 22%-34%) before the pandemic^4^, the prevalence of turnover intention among all nurses (18 studies) rises to 38% (95% CI 26%-51%) during the COVID-19 pandemic^5^. Also, the prevalence of intention to leave the profession among nurses (16 studies) reaches 28% (95% CI 21%-34%)^5^. One may argued that the high heterogeneity of studies included in meta-analysis and the adaptive strategies to promote nurses retention in post-pandemic era^6,7^ may lead to the over-estimation of rate of nurse turnover behavior though the turnover intention was regarded a good predictive indicator of actual behavior^8,9^. The coronavirus is still changing and killing when the new global alerts of infectious disease of devastating potential has been issued recently^10,11^. The post-shock health system may have not recovered completely from last pandemic let alone has been prepared for next wave of flooding in inpatients. The challenge of front-line medical workforce retention especially registered nurses and licensed doctors becomes even more serious and urgent.

Frequently, the studies on nurse turnover surveyed the prevalence of turnover intention and explored its risk factors or predictive factors^5,12–18^. These predictive factors can be roughly categorized into six themes in descending order hierarchically as organizational culture(support, justice, development opportunity), work conditions(resources, staffing), job demands(work load, time, pattern and complexity), employment services(experience, skills), work relationship(job control, reward, value, fairness, security) and personal characteristics(family, health)^5,19–21^. These postulated predictive factors of turnover intention may be helpful for policy makers at institutional level (local) in short-term. The cross-sectional surveys will neglect the problems in temporal scale and the heterogeneous surveys may prevent researchers to piece evidence together. As a result, the studies on nurse turnover intention has not reached a consensus conclusion that may be employed in further decision. Another setback of current nurse turnover studies is to use the turnover intention largely for convenience rather than depending on real turnover data while the actual turnover occurs frequently^22–25^.

We wonder whether the nurse turnover rate could be wound down at an individual institutional by adoption proposals provided in published papers when job opportunities would be available easily for registered nurse. A more broad background than the institutional level should be revealed and also a longitudinal perspective study should be provided^24,26^. By delving into the Chinese national statistics we noticed a rapid medical workforce growth in last two decades. During this period the growth of registered nurse exceeded that of the licensed doctors and becoming the largest workforce in health sector. This descriptive analysis of national statistics also revealed discrepancies of registered nurses in three tiers of hospitals which may be helpful to understand the current problems of nurse turnover and shortage in China.

## 2 Material and methods

### 2.1 Data

The annual statistics of registered nurse, hospital beds and other related data from public accessible website of the National Health Commission of People’s Republic of China^27^ (accessed on 2024-05-20). Twelve electronic version books (in Chinese) of Annual Statistics of China Health System and Status were available from 2011 t0 2022. In general, these statistics yearbooks provided fourteen categories of information covering the health institutions and beds, health personnel(including medical students), health hardware infrastructure, investment and income of health sector, medical service and patients visit, grass-roots health service, health serviced through Chinese traditional medicine, maternal and children health service(covering pregnancy to pre-school children), selected indices of population health, public health service(disease prevention and control), causes of death, food safety and its administration, medical insurance, demographic information and appendices some socio-economic indices. The definition of provided statistics also available and presented in supplement materials(English definiction of statistics). A brochure with bilingual glossaries, from year 2011 to 2019, were also provided at the same website, therefore, and we referred to these glossaries in Chinese to English translation. The nurse number data were also collected from other website (https://ourworldindata.org) as double check to correct any typos.

### 2.2 Statistical analysis

All data were used as in their original form, except we times the raw data accordingly when they were with unit of kilo- or higher magnitude, and no data clean up method has been used. The linear interpolation was used in estimate the elder population(≥65 year of age) among three population census data in 2000, 2010 and 2020. The data was manipulated and analyzed by using R(version 4.2.2).

## 3 Results

### 3.1 The rapid expansion of hospital beds corresponding to aging population in past two decades

All beds available for patients were 2.18 million in China in year 1980, thereafter, the bed number stumbled to reach 3.14 million in year 2002, which is the second year (first year in 1996) with bed number decreased in this period. Roughly, one million beds increased in twenty years, on average, 43.2 kilo-beds increase each year. Even though, the bed number in year 2003 (3.16 million) is less than that in year 2000 (3.18 million). After 2003, the bed number increased rapidly with 99.1-564.9 kilo-beds each year until 2021. As a result the total bed number peaked at 9.45 million in 2021, about three times of that in 2002, six million beds increased also in about twenty years (Figure 1A). This wave of beds expansion seems in coincidence with the growing population of the elderly in China(Figure 1B). In this period the proportion of bed number in all hospitals made up >70%, and peaked at >78% in 2020, while the proportion of bed number in general hospitals lingered between 43.1% and 49.7%, peaked at 54.4% in 2005 (Figure 1C). Two inflection points of general hospital bed numbers were noticeable in 2005 and 2015, respectively. The gap between all hospital-based beds and those from general hospital displayed a trend of widening, especially after 2015. These data suggested that hospitalized treatment had been highlighted in past two decades while the general hospital-based pattern remains relative conservative. The rapid expansion of beds available for patients may be view as a response to increased population of the elderly, in turn it also may have been the driving force of the increase of health professionals (e.g., doctor, nurse, therapist, pharmacist, laboratory and medical imaging technician etc) in the same period (Figure 1A).

**Figure 1.**
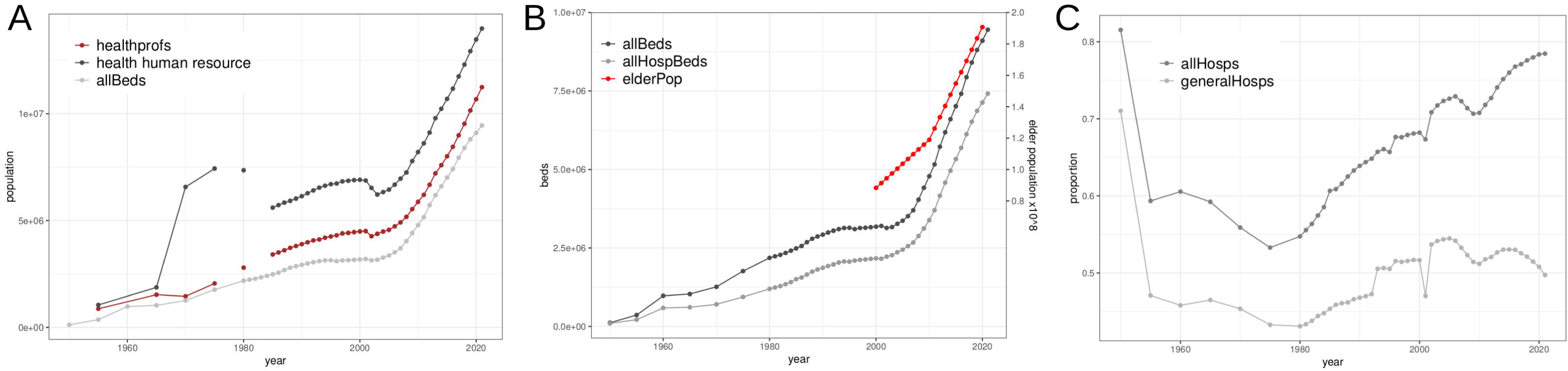
Beds and health professionals increase in response to aging population. The ratio of health professionals remains nearly constant proportion among all human resource in health sector from 2003 to 2021, both grows rapidly in parallel with the expansion of beds available(A); beds number grows faster than hospital-based beds though the latter still dominates(B); among hospital-based beds a large portion beds other than general hospital has appeared(C).

### 3.2 Registered nurse become the largest workforce in health system in last decade

At first, we asked a question whether the register nurses in shortage was a long-term and stubborn social problem. The annual increase of the registered nurse usually lagged behind those of licensed doctors (including licensed assistant doctors or stated otherwise) before 2005, when the annual increase of registered nurses over-numbered that of licensed doctors. The registered nurse population in health system did not over-numbered those of licensed doctors until year of 2014 (Figure 2A). It seems that the increase of registered nurses take the lead in the growth of health professionals even taking the whole population or health professionals or human resource in health sector into account (Figure 2B, 2C, sFigure 1A). A noticeable trend started in 2003, when the proportion of registered nurses in all health professionals grew steadily from 28.9% to 44.6% in 2021, while the proportion of licensed doctors in all health professionals decreased from 44.7% in 2005 to 38.1% in 2021 (Figure 2C). Change had happened, the registered nurses were now the leading workforce in health system in terms of population. However, this pattern of crossed growth in licensed doctors and registered nurse should be interpreted with cautions as the ratio of doctor to nurse changed rapidly from 1:0.65 in 2003 to 1:1.17 in 2021. If we took beds number into account, this crossed growth mode remains (Figure 2D). Considering the rapid expansion of beds in hospitals it is predictable the patient-caring work should be shouldered largely on the whole population of registered nurse while each doctor’s workload will be increased as the bed number per doctor increased if we assumed the clinical workload of doctors remains constant and can not transfer to others (including machines). If that were true the increased doctor to nurse ratio would turn out to be the problematic results of rapid beds expansion and the neglect of maintaining a suitable doctor-to-nurse rate in health service system accordingly.

**Figure 2.**
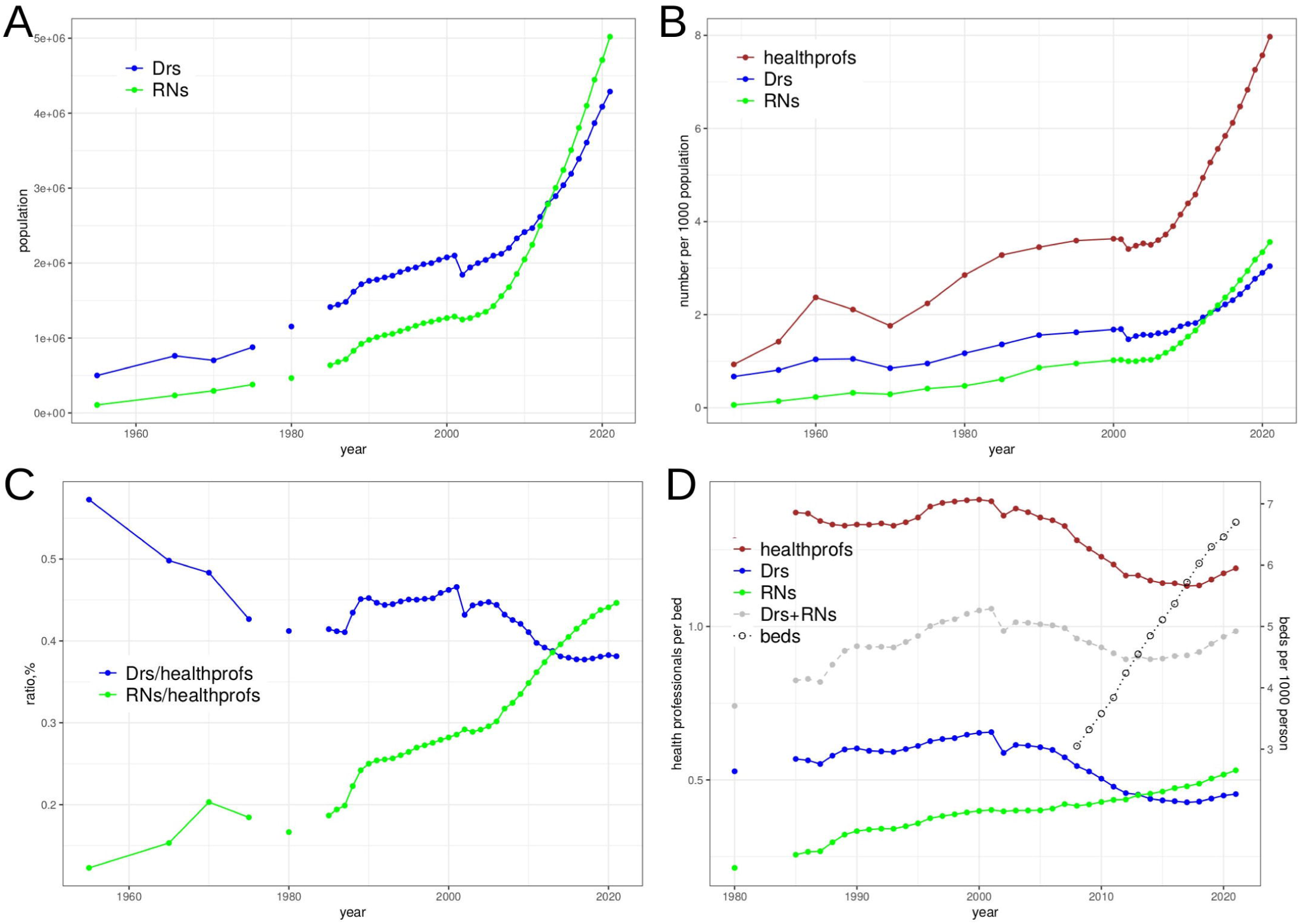
The registered nurse are leading workforce in health sector. In past decades the registered nurse population lags behind the licensed doctor population until last eight years(A); the registered nurses grows in parallel with all health professionals with respect to population since 2003 (B); in consistent with the absolute number the proportion of registered nurses to health professional elevated also (C); with the rapid beds expansion the growth of health professionals indicates slightly lowered proportion, even if that of the doctors, the growth of registered nurses, however, reverses this trend (D). The proportion of both licensed (assistant) doctors and registered nurses among all health professionals grew from less than half to about 80%; beds per 1000 person increased rapidly (near linear) in past decade, the rapid expansion of beds coincide with the lowering of doctors per bed (D).

### 3.3 A considerable proportion of medical graduates not enter health sector

If the rapid beds expansion had been planned in advance, the medical student recruitment and working opportunities provided for medical graduates would be planned in advance too. This is known as top-down design, still a preferred national strategy in China. Even a stunning number of registered nurses had supplied health sector, we still wonder whether medical education system failed to meet the human resource requirement of health system yet. The annual medical graduates from high- and mid-level education does increased rapidly. The medical graduates from high-education increased from 1.2×10^5^ in 2003 to 9.4×10^5^ in 2021, the proportion of medical graduates in all graduates of high-education grew from 6.2% to 10.4% in the same period. The averaged increment of high-education medical graduates were 43894 persons each year, and were 55747 taking mid-education medical graduates into account (Figure 3A). In last six years the high-education medical graduates over-numbered the mid-education medical graduates with a rate between 1.25-2.14:1. However, the recruitment of health professionals in health sector seemed staggered and reluctant, as a result the gap between the annual increment of health professionals and medical graduates became wider (Figure 3A). The annual increment of health professionals approximately equal the new recruitment of medical graduates minus the retired medical professionals, and the latter number remained unknown.

**Figure 3.**
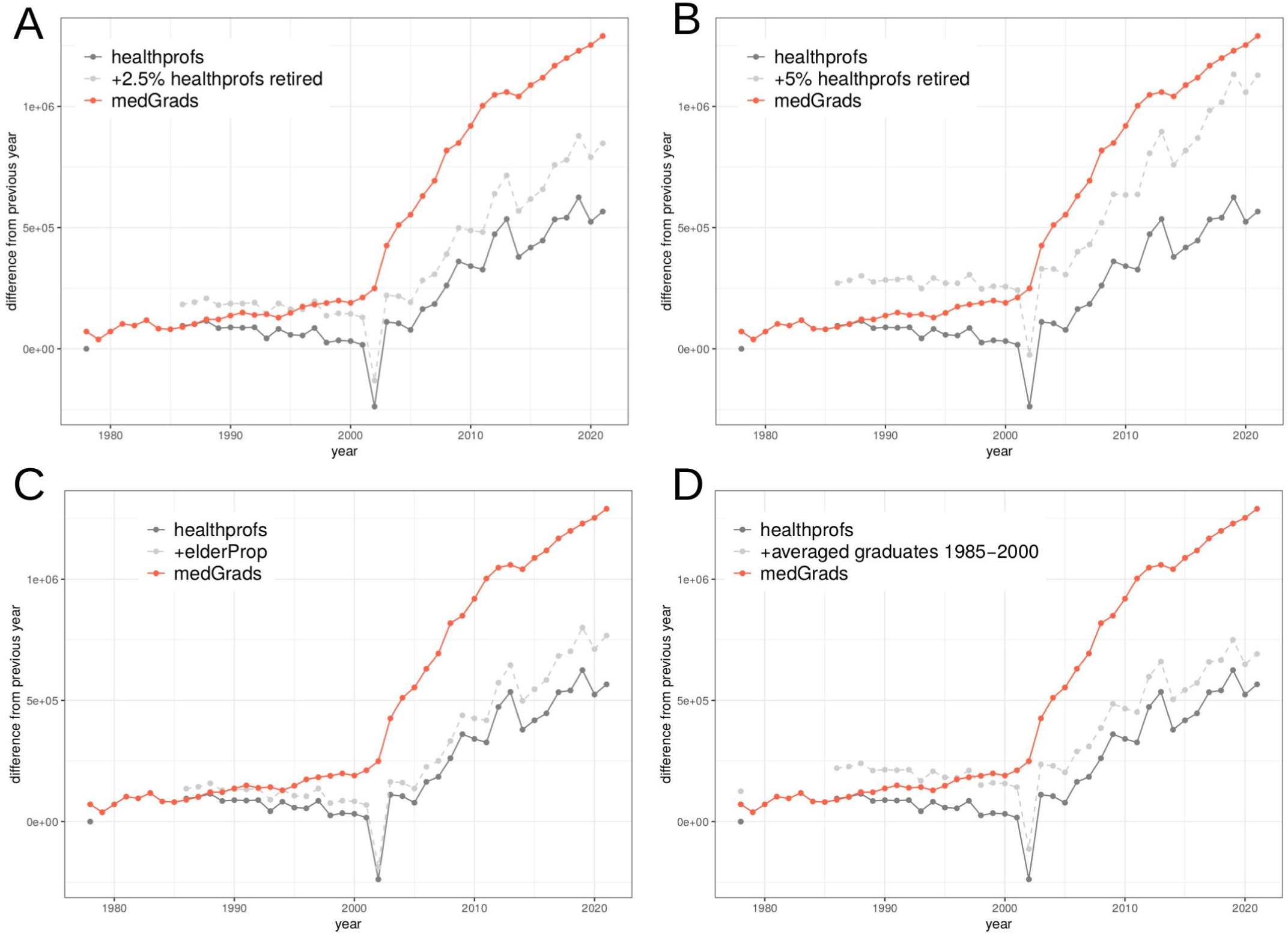
Estimation of medical graduates entering into health sector. Both the annual increment of health professionals and medical graduates (including high- and mid-level education) were accelerated since 2003. When retirement replacement was estimated by using 2.5% (A) and 5% (B) of all health professionals; and proportion of elder population (C) and averaged annual graduates from 1985 to 2000 years (D). Batch retirement from their 25-60 years work at 2020(n=125113). **Notes:** the proportion of 60 years of age among working age population of 25-60 years was used as elderly proportion in this case.

Therefore, several methods were used to estimate the number of retired health professionals in each year. (Method01)A proportion of population at 50 years would retire at their 60 years in next decade. By using the population census data in 2000 and 2010, the people in their 40 years and 50 years, which were anticipated to retire at year of 2020, were about 1.89% and 2.24%, respectively(sFig 2). A proportion of 2.5% of all health professionals was used to estimate the retired health professionals still indicate gap between the new recruitment and medical graduates became wider gradually (Figure 3A). This trend changed when 5% of all health professionals as used to estimate the retired health professionals (Figure 3B). As the elder population growth the proportion of retired health professionals may grow accordingly. (Method02)A retired proportion of health professionals were estimated by using estimated proportion of the elderly(age of 60 year in this case) in each year (by using interpolation between 2010 and 2020). The widening gap between the new recruitment of health professionals and medical graduates retained (Figure 3C). (Method03)This trend remained even we used the averaged medical graduates number from 1985 to 2000 (125113 person), they would retire after about 35 years of work (in 2020) (Figure 3D).

Given a proportion of 2.5% of all health professionals would retire each year a considerable proportion of medical graduates (including high- and mid-education) might fail to enter health sector, these proportions ranged from 28.5% in 2019 to 65.3% in 2005, averaged 44.5% between 2003 and 2021 (Figure 3A).

No matter how to interpret this large portion of medical graduates not entering the health sector each year in past two decades, this portion of medical graduates would not contribute to the duty of care of hospitalized patients (estimated based upon all hospital-based beds available). Therefore, it seemed that the medical student enrollment would have been planned in advance, but the health sector seemed only accept about 3/5 of them.

### 3.4 The workload discrepancy of registered nurse in three tiers of hospitals

Despite the registered nurse became the leading workforce in health sector in terms of population, still a considerable proportion of medical graduates can not contribute to hospital bedside care though the actual number of nurses among them was unknown. We then considered whether the nurses in hospitals, where their professional nursing skills were in urgent need, had been distributed and managed optimally. Greater than 90% of hospital-visit-patients, ranged from 90.6% to 97.9% from year 1985 to 2021, visited outpatient and emergency department. The proportion of hospital admission grew steadily from 2.1%(1987) to 5.7%(2020) among all hospital-visit-patients, from 29.26 million to 183.52 million in the same period.

Fortunately, the year 2003 was the inflection point once again, the patients’ visit to hospital increased rapidly thereafter (Figure 4A). However, three tiers of hospitals responded differently though, on the whole, they all took more inpatients each year in past decade. When the annual inpatients number in tier 1 hospitals grew slowly, from 4.6 million to 11.2 million, the annual inpatients number in tier 3 hospitals grew from 6.6 fold to 10.0 fold of those in tier 1 hospitals, from 30.96 million (2010) to 112.52 million (2021). On contrast, the growth speed of annual inpatients number in tier 2 hospitals decreased from 11.0 fold to 6.1 fold of those in tier 1 hospitals, a cross of fold trajectories occurred in 2016 (Figure 4B, 4C). Four years ago the number of registered nurses in tier 3 hospitals surpassed that in tier 2 hospitals(Figure 4D). The following surge of inpatients in tier 3 hospital may attribute partially to increased registered nurses, increased capacity to care more inpatients. If we considered the annual workload in terms of inpatient number per nurse, the tier 3 hospitals was lower than that in tier 2 hospitals until 2020, even lower than that in tier 1 hospitals before 2017 (Figure 4E). Also we can read from the data that the ambitious expansion of tier 3 hospital beds were prepared themselves for attracting more inpatients, this trend also triggered the changes.

**Figure 4.**
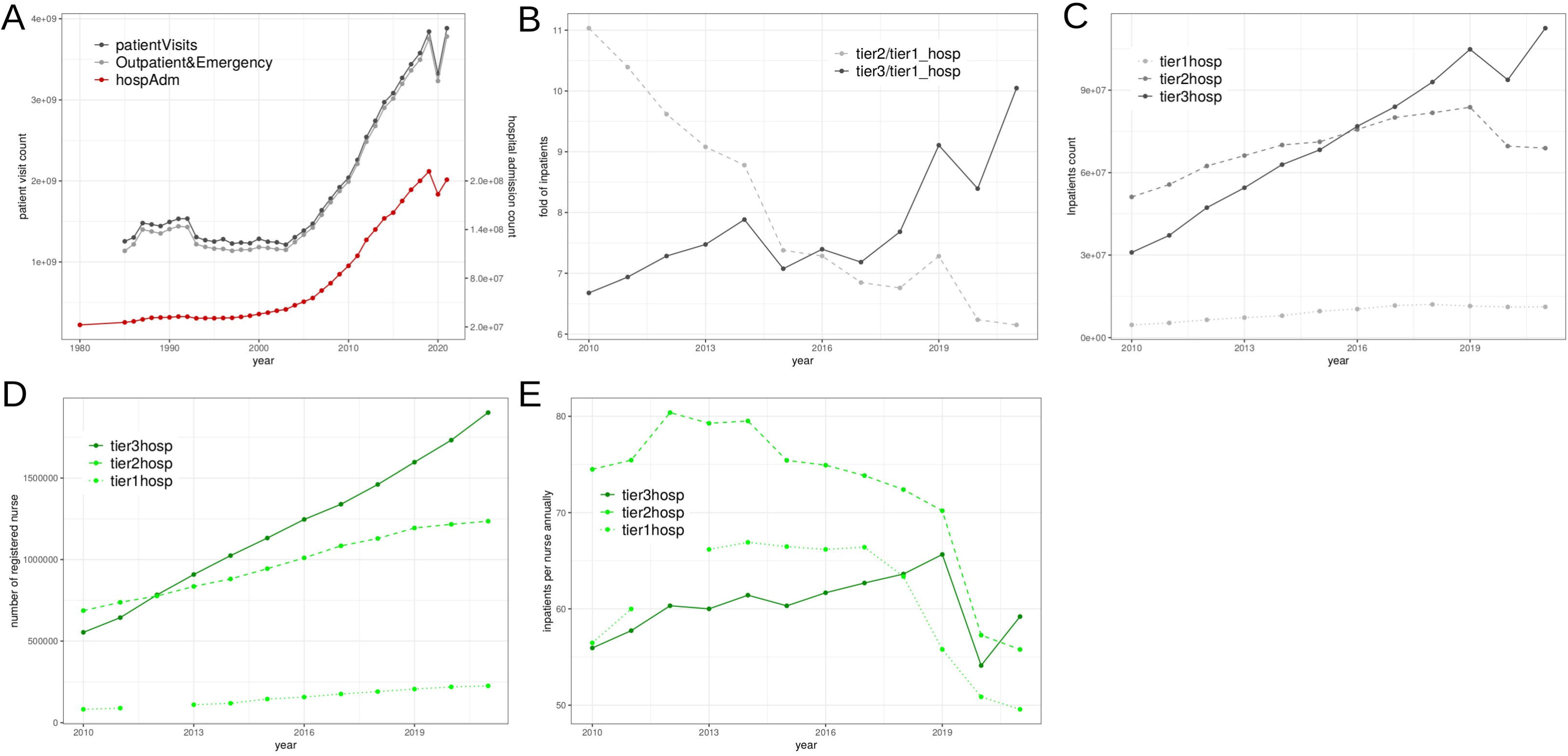
Inpatient and inpatient-to-nurse. Annual patients visited hospitals almost through outpatient and emergency department, the hospital admission rate kept nearly constant(A); the inpatient number presented as fold of that of tier 1 hospitals displayed a cross-over at 2016 (B); the actual inpatients in tier 2 and tier 3 hospitals indicated the cross-over at same year, inpatients number rebounded back only in tier 3 hospitals during pandemic (C); the registered nurses increased more rapidly in tier 3 hospitals and over-numbered those in tier 2 hospitals around four years ahead of the over-number of inpatients (D); with respect to inpatient number per nurse the tier 2 hospitals took the lead for a decade before pandemic (E).

### 3.5 The nursing care income discrepancy in three tiers of hospitals

We employed other indices to see whether the roughly estimation of inpatient number per nurse as workload in three tiers of hospitals would be misleading. The hospital level nursing income indicates the chargeable service of nursing care or direct operations in hospitals. From 2012 to 2021 the nursing care income of tier 3 hospitals was 3.9-5.2 folds that of tier 2 hospitals, and was 39.5-55.0 folds that of tier 1 hospitals (Figure 5A). The nursing care income per nurse of tier 3 hospitals was 3.3-4.4 folds that of the tier 2 hospitals, and was 4.8-6.8 folds that of the tier 1 hospitals. The nursing care income per bed of tier 3 hospitals was 4.2-5.5 folds that of tier 2 hospitals, and was 8.2-12.4 folds that of tier 1 hospitals (Figure 5B,C). After normalized the nursing care income by using the number of hospital beds, nurses and inpatients (merely on purpose of comparison), the nursing care income of tier 3 hospitals was 13.2%-22.9% of that of the tier 1 hospitals, and this rate was 2.6%-8.3% for tier 2 hospitals (Figure 5D). The nursing care income was largely diluted by bed number, this data suggested the expansion of hospital beds may have reached its limit of safe. These data indicated that the nursing care income in tier 1 hospitals was more labor-intensive than those in tier 2 and tier 3 hospitals. In other words, the nursing care for each patient, each bed, each nurse would be more human-dependent in tier 1 hospitals though it was on sharp decline from 2013 to 2015. Each inpatient in tier 1 hospital would receive more professional care directly from nurse than that in tier 2 and tier 3 hospital from 2012 to 2021(in terms of nursing care income). Another interesting finding was the income of bed rent (charge for bed fee by day in hospital) was higher than nursing care income in all three tiers of hospitals (Figure 5A). In consistent with these results the rate of nursing care income to the income of hospitalized patients in tier 1 hospitals was the highest among three tiers of hospitals and all were on the rise, except the places of tier 2 and tier 3 hospitals exchanged. If the growing nursing care income could be viewed as direct result of chargeable nursing service it might be attributed partly to increased nurse workload and to the enlarged nurse population. Accordingly, the number of beds per nurse in tier 1 hospitals was highest among three tiers of hospitals (sFig 3C,3D). The nurses in tier 1 hospitals need to do more nursing care work for each inpatients, while the nurses in tier 2 and tier 3 hospitals need to care much more inpatients. The workload may increase for all registered nurses but from different sources.

**Figure 5.**
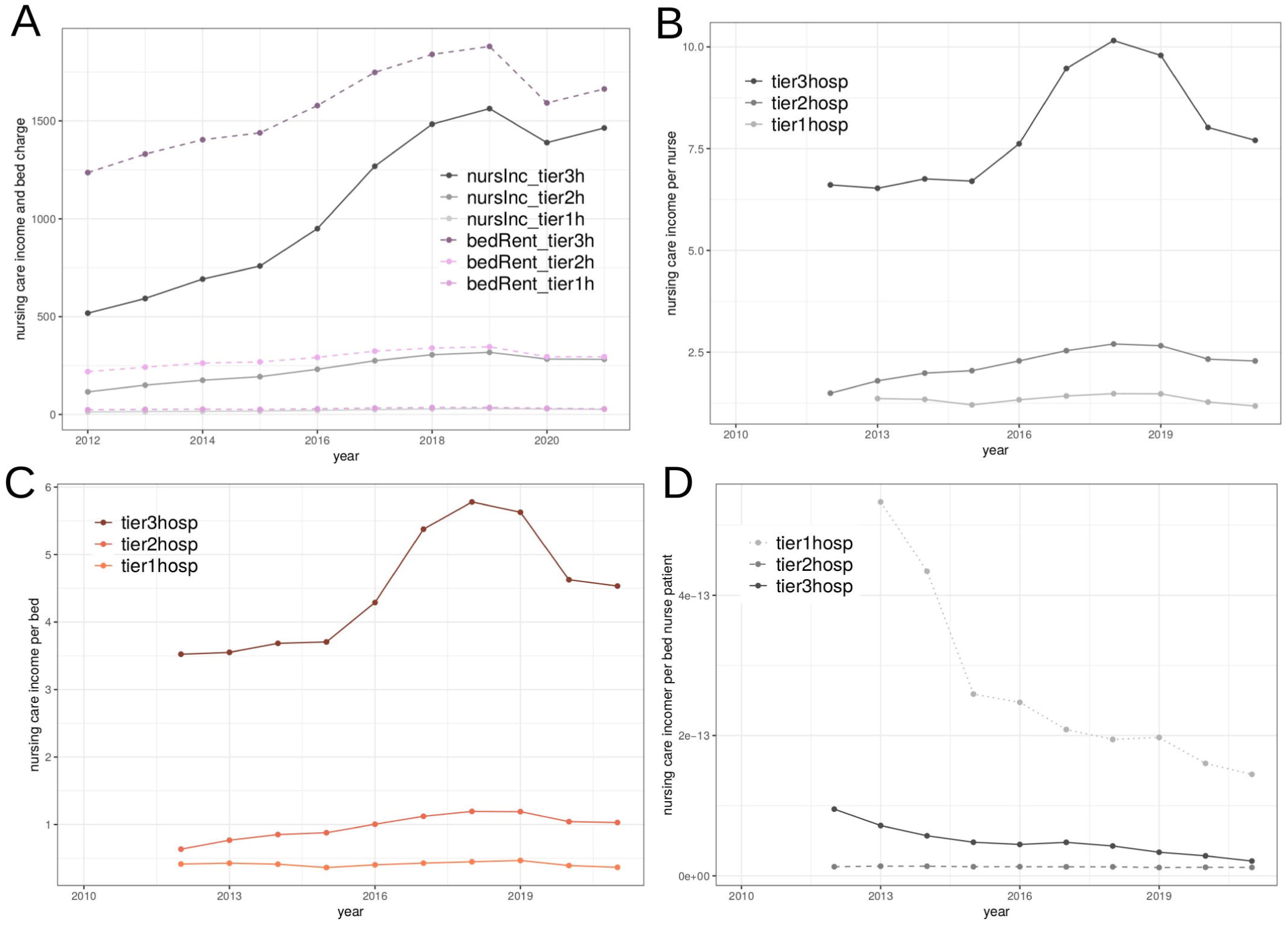
Nursing care income in three tiers of hospitals. The nursing care income was less than bed rent (in 10^4 CNY) in all three tiers of hospital though the gap (especially in tier 2 and 3 hospitals) was closing in past decade, the higher in tier the larger of income gap between bed rent and nursing care income(A); the nursing care income per bed or per nurse surged once in tier 3 hospitals from 2015-2018 coincide with bed occupancy rate decrease but reasons is unclear(B, C); in terms of nursing care income per bed, per nurse and per inpatient, tier 1 hospitals is highest though all were decreasing(D).

### 3.6 The city and rural discrepancy in nursing workload

Nursing care income in tier 1 hospitals was more human labor-intensive because all tier 1 hospitals were less equipped with instruments and medical professionals as hierarchical design of three tiers of hospitals in many dimensions for different purposes. Greater than half of hospital beds were deployed in vast rural areas from 2010 to 2020, this trend shifted in 2021 as a result of rapid city expansion and large volume of rural-to-city immigration. The number of registered nurse was greater in city than in rural area while the number of beds per nurse in rural was higher than that in the city (Figure 6A,B). Currently we lack data of inpatients per bed of city and rural, it is quite possible that the inpatients per bed in city will over-number that of in rural. This city and rural nursing workload discrepancy had been existed for at least ten years though the gap is closing.

**Figure 6.**
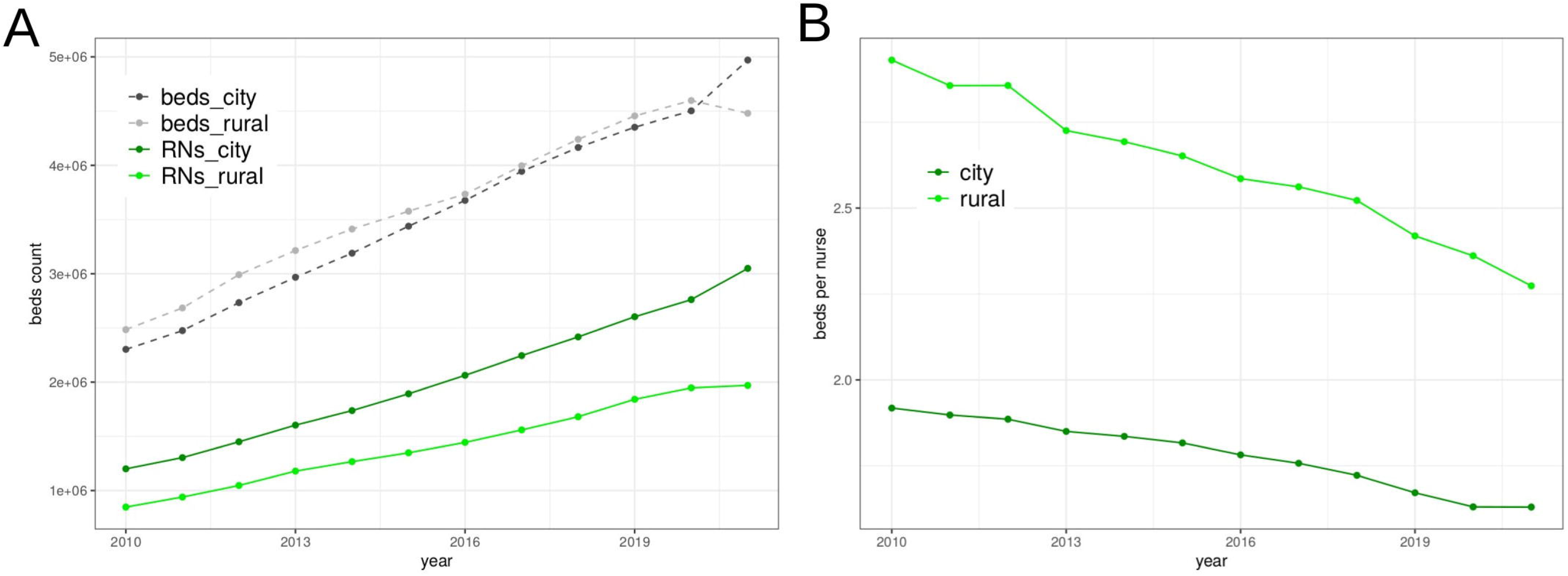
City and rural discrepancy. In total, the vast rural area boast more beds than city downtown but nurse population in city is greater than that in rural(A); estimated beds per nurse is lower in city than in rural area (B).

## 4 Discussion

The overall background is the Chinese population is experiencing rapid aging in past two decades and yet the coming two decades. During the first half we expanded the hospital beds hurriedly in response to this wave of aging, however, the looming ride is even more serious (Figure 1B, sFig 2). The majority of hospital beds and registered nurses had been largely distributed in tier 3 hospitals, lead to the overwhelmed workload for both doctors and nurses accordingly. As a result the doctors and nurses population indicated a high rate of turnover (ranged from 5% to >60%) ^28–32^ and burnout^21,33–40^. The discrepancy in hospital beds expansion in three tiers of hospitals, especially in last decade, has lead to changes, however, the inherent discrepancies retained largely, also was the city and rural discrepancy. The flooding in inpatients and corresponding increased nursing workforce may enlarge the existing discrepancies in three tiers of hospitals whatever the exact reasons for hospital beds expansion and distribution might be (Figure 4, 5). This study provided a profile of registered nurse population in health system in last two decades to help understand the high rate of turnover and burnout, which may cause serious subsequences^41,42^; and to minimize the improper interpretation of statistics, which may lead to further misunderstanding and confusions.

It is not surprising that year 2003 became a pivotal followed by near two decades of rapid hospital beds expansion, because China seen an unexpected outbreak of SARS-associated coronavirus early in that year^43^. That epidemic might have been an acid test for health system in China. The national statistics displayed that the hospital beds have been largely deployed in hospitals of tier 3 level, which were leading in instruments and medical professionals among three tiers of hospitals. Some times the tier 2 hospitals could upgrade to tier 3 when the criteria were met, however, the criteria would vary. This fast track to build tier 3 hospitals can be viewed as active response to rapid aging Chinese population. Whether we should prepare more hospitalized beds for all inpatients or we should treat patients selectively (not defined yet) remains unknown. It should be cautious as the hospital beds increased constantly at the cost of elevated debt rate, from about 30% to 40% in 2012-2020, while the rate of bed occupancy decreased gradually in the same period(sFig 4A). The registered nurses are direct nursing care provider, the nurse staffing and quality of care are critical for outcomes and prognoses of inpatients^44–53^. The health sector were usually estimated to take in 70% of medical graduates^1^. If 2.5% of health professionals would retire and be replaced by newly graduates each year, the increment of health professionals was about 56% of the new graduates in the same year. This rate was about 75% of the new graduates if 5.0% of health professionals would be replaced each year(half of health professionals would retire in following ten years). In 2021, the registered nurse made up of 44.6% of health professionals in 2021(still lower than global level of 59%); the increment of registered nurse was 310705, made up of 24.1% of all medical graduates, 32.9% and 89.6% of high- and mid-level of medical education graduates in the same year, respectively. The medical graduates were increased almost constantly in last two decades while the increment of registered nurses was fluctuated in the same period, especially in year of 2002, 2008, 2014 and 2020 (sFig 4B). On the other hand, the overall loss of medical professionals also including those quit their professionals prematurely. The exact number of these population was unknown. In this study we use only the estimation of proportion of retirement of medical professionals, a gap between the data including only retired medical professionals and actual reception of medical graduates will be a rough estimation of loss of medical professionals prematurely, no matter whether they are failed in transition from theory to practice in early stage or attrition due to other reasons latter^54–58^. A question on the quality of nurse graduates was risen as the medical (education) system had to train such a constant increasing population of nurse students based upon shortages of (human/teacher) resources as well, if we presumed the experienced nurse teachers are also needed in proportion to students. If the qualified medical graduates did not increase in proportion to the increased medical graduates this means the (human) resources invested into medical education system had been wasted largely. The increase of licensed doctors was much slower than that of registered nurses and this can be viewed as a red sign for nurse education if we assumed the medical training of both doctors and nurses were similar(Figure 2A).

It would be confusing that the estimated nurse turnover rate was 0.75%-4.60% among thirty provinces by using data from 940 hospitals (3.0% of all hospitals in 2017) with complete records of nurse turnover (n=816656)^59^. From 2008 to 2018, four other surveys (n=4320 in total) indicated nurse turnover rate of 19.6%(95% CI, 10.8%, 35.5%) in China, quite close to the synthesized results of global prevalence 18% (95% CI, 11%-26%)^20^. This rate is still about 10% lower than the global prevalence of turnover intention (both before COVID-19 pandemic)^4^. This study will be helpful to understand the different results from above mentioned surveys.

We used the inpatient number as a direct index of workload for registered nurses because most of them required bed-side nursing care. The patient-to-nurse ratio (or its reciprocal) has been used to indicate the risk of insufficient nurse staffing and has even considered in future legislation^60–63^. Moreover, the flooding in inpatients that might have overwhelmed the registered nurses easily was de facto in China. The long-term work burden overload strained the vulnerable health system and intensified the conflicts among them, the patient-oriented violence against medical professionals was not unusual and even at life cost^64–66^. A proper patient-to-nurse ratio may involve raised cost of human resource, however, this could not be the reason to cut salary for registered nurse as common seen in China hospitals^28,28,32,67–71^, whether it would be hospital-dependent was not merely an economic problem^61^. As for patient-to-nurse ratio should be job-dependent even in individual hospital, and the hospital should be empowered capable of decided by themselves.

We are surprised to learn that the bed rent is higher than nursing care income in three tiers of hospitals. At least the nursing care income indicates the skillful operations of qualified clinical nurses on inpatients.

Whether the bed rent was an economic lever to compensate for the low nursing care income or the nursing care income list had to take the least developed areas into account (same standard across China mainland) and, as a resulted to control it deliberately remained unknown. It would be natural to regard that the nursing care is cheap and so as their skills and operations, the nursing care skills are dispensable and the nurse staffing on the whole might have been regarded as a burden economically for a hospital. The imbalance of workload among three tiers of hospitals and distorted nursing care income can be viewed as two indicators of inherent structural problems of health system. It will be predictable if the nurse remuneration was determined merely by local economic conditions with reference to nursing care income, the mobility of registered nurse from low to high income area would be common even if high workload challenge. This study did not provide further data to explain the underlying reasons, however, these statistics triggered our curiosity.

We have seen the sharp decrease of inpatients due to areal lock down during the pandemic and inpatient number rebounded only in tier 3 hospitals (Figure 4A). It seems the high quality medical services attracted more inpatients. If tier 2 hospitals could upgrade to tier 3 hospitals with specified conditions, it seems that the tier 1 and tier 2 hospitals should provide high quality medical service to attract as more inpatients as possible to avoid to be marginalized in the future. It’s good thing to provide high quality health service for Chinese population. Currently, the discrepancies among three tiers of hospitals had displayed some trends, whether seemingly more job opportunities and challenges for registered nurses would arise the ambition in them and encouraged them into an elevated turnover rate remained unclear. Due to discrepancies whether the nurse turnover among three tiers of hospitals would be beneficial for spreading of experiences and skills required further study, especially when the newly graduated nurses are not qualified sufficiently. High rate of nurse turnover then was an adaptive and corollary result.

We wonder whether to increase number of nurse hurriedly can be a solution to over-numbered inpatients in population aging. When a large proportion of nurses failed to enter health sector, they have to leave their profession or be left behind in rural areas, the nursing care skills may have not been wasted completely, whether we should encourage and mobilize the people with professional nursing skills with systematic support requires further study. It seems that the analysis of national health statistics on registered nurses have opened an avenue to raise more questions than these data can lead to a tentative conclusion. From the missing data of statistics it becomes apparent that the city and rural should have been regarded and treated different naturally for a long period. Last but not least, who would care the care giver as few of the nurse-centered surveys concerns how to deal with difficulties from the point of view of nurses themselves. The problems would inevitably remain if how to provide organizational support and justice, how to improve working environment, how to upgrade the skills to deal with job demands, how to elevate the professional identification and workload-based remuneration and how to retained qualified nurses as much as possible were unsolved. It is a common sense that one can hardly expected high quality humanized care if the direct care giver were treated unfairly in a merciless environment all day long.

### Strengths

This study presented a longitudinal perspective of registered nurse population in past two decades, a nurse boomer in response to aging population with rapid beds expansion; used the national level statistics based upon the inquiry of general reasons underlying of nurse shortage and high rate of turnover; revealed the inherent structural problems of three tiers of hospitals and challenges of medical professionals in this system, discrepancy between city and rural. These information might be useful both for institution and individual nurse. We provided a panorama view to understand the shortage and high turnover rate of nurse in China though use only the statistics available.

### Limitations

some definitions of statistics have evolved from 2002 to 2020, the influence on statistics of health personnel was estimated less than 2.6% (358000/13985363) based upon data in 2021. Macro-scale data are results of contributions converged from all levels of health institutions/hospitals, therefore, the problems of such scale may only be solved by flexible, adaptive and reiterative improvement in multiple aspects. We don’t anticipate any one-for-all rules can solve macro-scale problem easily. One may argue that the national level statistics could be high heterogeneous due to the great differences among vast areas in China, to some extent we hope that the variations both positive and negative could be somewhat cancel out partially and the final statistics would be much close to the expected level on average.

## 5 Conclusion and interpretation

This descriptive analysis of national statistics of registered nurses in China revealed the positive response to aging population in past two decades with beds expansion and largely increment of medical professionals, especially registered nurses as to now the leading workforce. A large portion of medical students may have failed to enter health system. The distribution of beds predominantly in tier 3 hospitals may have attracted more patients and lead to large gap among three tiers of hospitals, the discrepancies in three tiers of hospitals or registered nurses were inherent systematic problems. The discrepancies among three tiers of hospitals and, city and rural areas have caused concerns.

## Supporting information

Supplement material_definition

Supplement_material_sFigs

## Data Availability

All raw data are available online at http://www.nhc.gov.cn/mohwsbwstjxxzx/tjzxtjsj/tjsj_list.shtml

## Acknowledgements

We are grateful for all people who have contributed to the national health statistics, from the smallest primary care health station to the largest referential medical centers, and those who have worked hard to compile and organize these data for publish. We thanks Cuifei Song for critical discussion of nursing care income.

## Credit of Author contribution

LY, LQ, GZ conceptualize and design the study, collect and analyzed data; LY and GZ drafts the manuscript; LY, LQ, GZ read, discuss and modify the manuscript.

## Data availability

All raw data are available online at http://www.nhc.gov.cn/mohwsbwstjxxzx/tjzxtjsj/tjsj_list.shtml.

## Supplement materials

The supplement materials of this article contains figures and English definitions of statistics.

