## Supplement material_definition for "Past and present of registered nurse in China--descriptive analysis of longitudinal national statistics of registered nurse"

**English definitions of statistics**

1 From 2002 onwards, the number of medical and health institutions is counted by the actual registered number in departments of health, business and civil affairs; and the number of medical and health institutions is counted by the number authorized to be set up by the health or other administrative departments in the period 1949-2001.

**Notes**: this may influence the number of beds count before and after 2002.

2 Health human resource included employees working in hospitals, primary health-care centers, public health institutions and other health-care institutions, namely, included health professionals, rural general practitioners(assistant) and entry-level medical technicians, other professionals, managers and health-care assistant in clinical care. In practice, the statistics of health human resource were counted based on on-the-job employees who were paid year-end salaries, including all types institutional tenure staffs (including contractual staff) and ex-employees invited back to work for more than half a year, but excluding temporary workers, retired staffs and those staffs only retain contractual relationship on paper.

**Notes**: the health professionals included those ex-employees invited back to work for more than half a year since 2007; and included those "health supervisor" as civil servants since 2010; and included those personnel of family planning service department since 2013.

3 Health professionals includes licensed (assistant) doctors, registered nurses, pharmacists, laboratory technicians, imaging technicians, health supervisors, and health professionals in their internship (pharmacists, nurses, technicians), but excluding health professionals who had engaged in management (e.g., deans, vice deans, secretaries of China Communist Party, etc.)

Notes: the dentist in hospital belongs to licensed doctors; the health professionals did not include technicians in pharmacy or in laboratory (not graduated from medical education system) since 2007; the licensed (assistant) doctors only included those who have obtained their certificate of licensed (assistant) doctors from 2002 on; before 2002, the licensed (assistant) doctors statistics were based upon those on job. The licensed (assistant) doctors included those working in village health service stations. The licensed (assistant) doctors included four categories: clinical medicine, traditional Chinese medicine, dental medicine, and preventive medicine(public health).

4 Registered nurses are those who have obtained a certificate of registered nurse and is actually engaged in nursing job, excluding those nurses engaged in management.

**Notes**: statistics were based on the number of registered nurses from 2002 on, and based upon the actual number of nurses on job before 2002.

5 barefoot doctor(1968-09 to 1985-01);

Country doctor (village doctor) are those defined only by working place in countryside, including entry-level technicians, mid-level medical technicians (licensed assistant doctor) and licensed doctors (including general practitioners), both of medicine or Chinese traditional medicine.

**Notes**: the statistics of health system and status yearbook has categorize those health institutions in the county level or lower as grass-root health institutions but overlapped with the criteria of three tiers of hospitals in tier 1 and tier 2 hospitals distributed in county or countryside. The statistics of grass-root health institutions was not used in this study.
