## Supplement_material_sFigs for "Past and present of registered nurse in China--descriptive analysis of longitudinal national statistics of registered nurse"

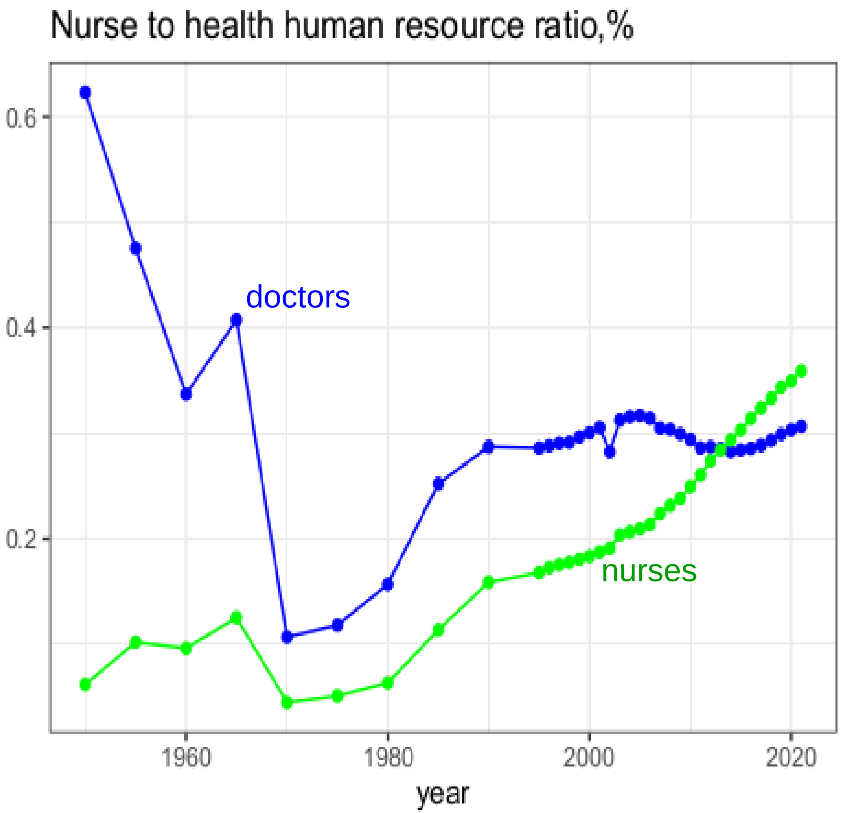


**sFig 1** Registered nurse and licensed doctors to all human resource in health sector

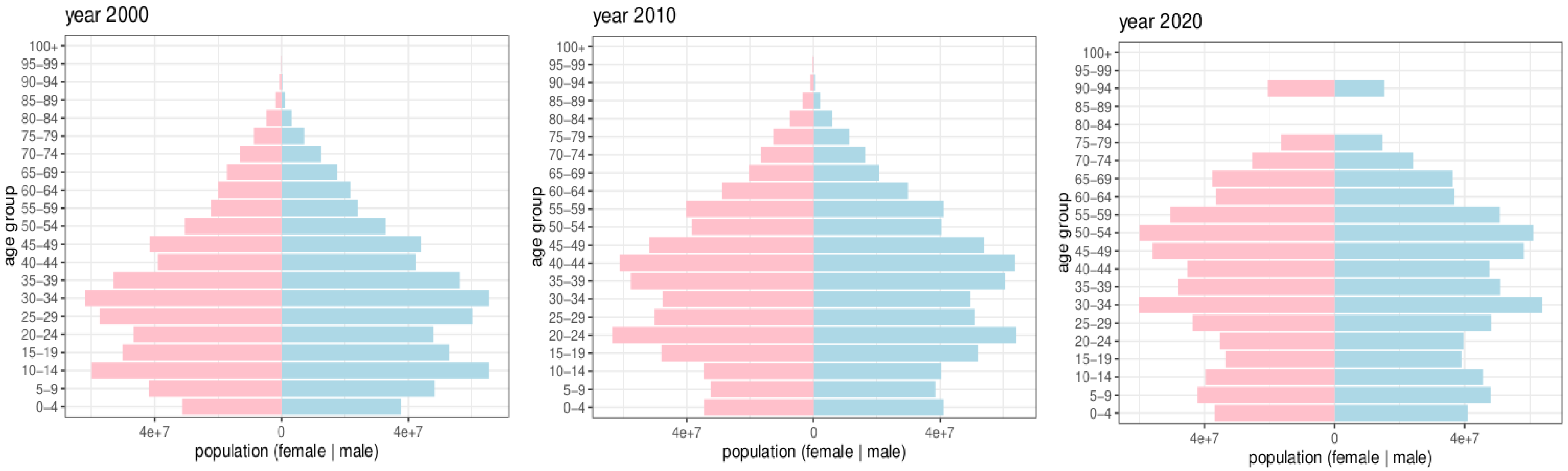


**sFig 2** Population pyramid by using census data in 2000, 2010, 2020

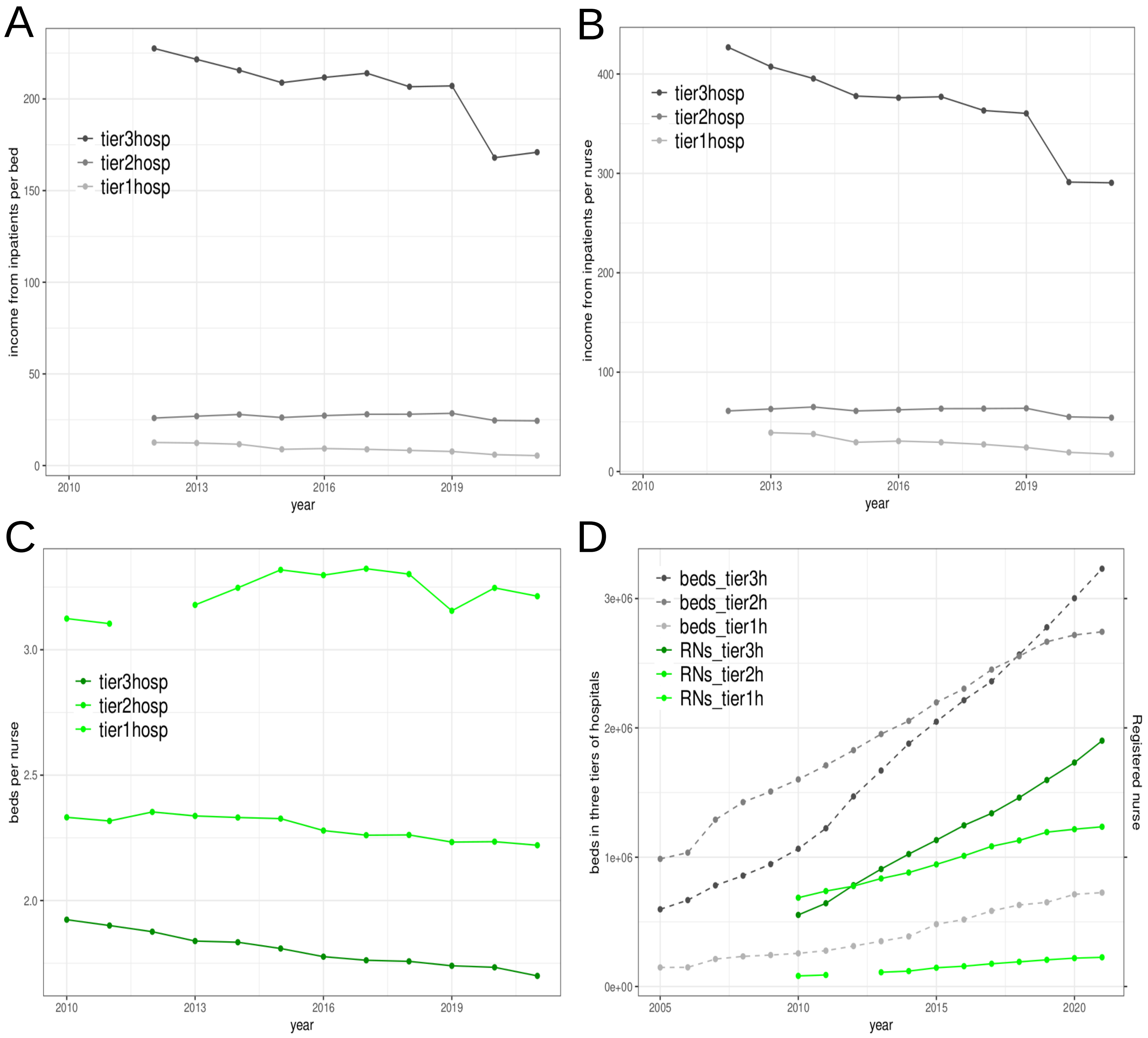


**sFig 3** The rate of nursing care income to income from inpatients(A,B,C,D)

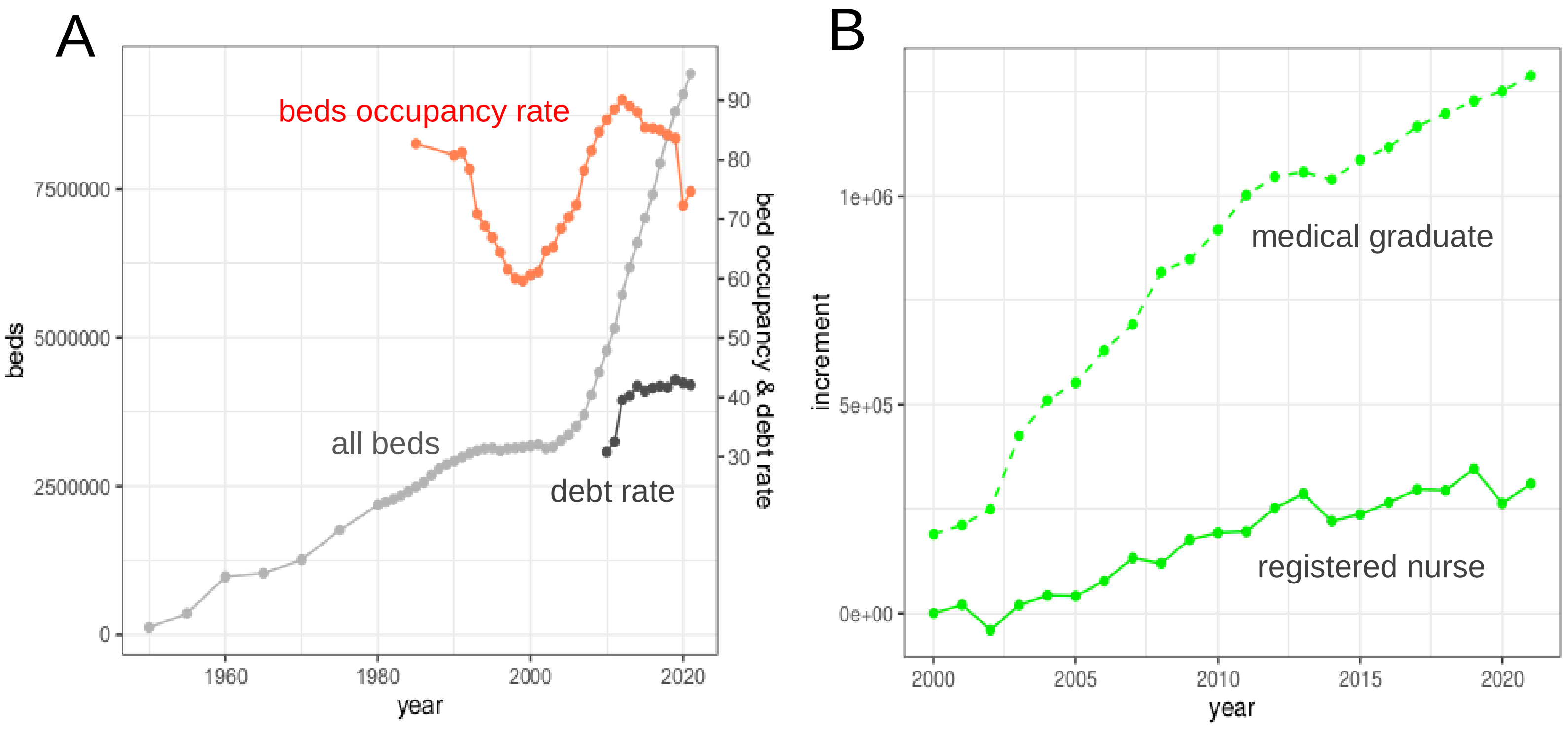


**sFig 4** Hospital beds expansion and bed occupancy rate and debt rate (A); the linear beds expansion in last two decades was not accompanied by constant bed occupancy rate growth but peaked at around 2010 and thereafter lowered gradually; meanwhile, the debt rate in all property increased from around 30% to 40%. It seemed that the bed expansion over-numbered the necessity and might risk in elevated burden of debt. Medical graduates (including high- and mid-level education) and yearly increment of registered nurse, the gap is widening (B);
